# An evaluation of the introduction of telehealth for remote antenatal and postnatal contacts in Bangladesh and Lao People’s Democratic Republic during the COVID-19 pandemic

**DOI:** 10.1101/2022.06.24.22276872

**Authors:** Sabera Turkmani, Rachel M Smith, Annie Tan, Catherine Breen Kamkong, Rondi Anderson, Siriphone Sakulku, Tej Ram Jat, Animesh Biswas, Caroline SE Homer

## Abstract

From 2020, COVID-19 spread rapidly around the globe and continues to have a major impact on health system functioning, with a disproportionate impact on low- and middle-income countries (LMIC). Reduced service utilisation and coverage of essential childbirth interventions is likely impacting maternal and newborn morbidity and mortality. Telehealth has been identified as an important tool in the continued provision of essential health care services. The aim of this study was to explore the experience and impact of implementing telehealth services for the provision of remote antenatal (ANC) and postnatal (PNC) contacts in regions of Bangladesh and Lao People’s Democratic Republic through 100 semi-structured interviews with health service leaders and providers, and childbearing women who organised, provided, or were the recipients of ANC and PNC telehealth during the COVID-19 pandemic response. The findings showed that a sudden pivot from face-to-face to telehealth services posed health system and care was challenging. Health systems lacked funding to support telehealth and the infrastructure needed for service changes; however, some were able to work with key maternal child health departments within Ministries of Health to find the resources to implement the services. Health providers found telehealth beneficial during the pandemic response but identified a lack of training, guidance, and support as a barrier to changing practice. Childbearing women reported being fearful of accessing care at health services due to COVID-19, and whilst they appreciated the telehealth contacts, many continued to prefer face-to-face delivery of ANC and PNC care. Telehealth, however, was a good alternative in a time when face-to-face care was not possible. Considerations for post-pandemic broader implementation or scale-up of telehealth for routine ante natal and post natal maternity care provision include the need for further research on issues such as accessibility, acceptability, quality of care, and sustainability of service provision.

## INTRODUCTION

COVID-19 infections spread rapidly around the globe and continue to have a major impact on health system functioning, with a disproportionate impact on low- and middle-income countries (LMIC) (1, 2). Whilst the direct impact on health systems is evident, the indirect impact of the pandemic on population health and wellbeing is far-reaching (3). Public health emergencies stretch even the most well-resourced health systems. Health care workers on the front line often feel the brunt of the emergency response. Long hours, irregular shifts and working outside the usual scope of practice are common experiences for healthcare workers during an emergency (4). These experiences are not limited to acute-care services, but impact primary service provision and this includes maternity services and in particular, routine antenatal (ANC) and postnatal (PNC) care as, unlike labour and birth, these may be seen as non-urgent.

COVID-19 has directly impacted maternal and newborn health. Pregnant women are at increased risk of experiencing severe illness when compared with the non-pregnant population. In addition to severe maternal illness, COVID-19 doubles the risk of stillbirth and may be associated with an increased risk for a small for gestational age and preterm birth (5).The indirect impact of COVID-19 on maternal and newborn health has also been substantial. Modelling estimated that in the least severe global scenario, there are likely to be 253,500 additional child deaths and 12,200 additional maternal deaths (3). The most severe scenario will result in 1157,000 additional child deaths and 56,700 additional maternal deaths. The reduced coverage of four childbirth interventions (parenteral administration of uterotonics, antibiotics, and anticonvulsants, and clean birth environments) would account for approximately 60% of additional maternal deaths (3).

In addition to the reduced coverage of essential childbirth interventions, health services and public health responses to COVID 19 led to disruption of routine healthcare services (5-7). In many countries, diversion of resources and a shift of focus from preventative health to acute management of COVID-19 de-prioritised sexual, reproductive, maternal, newborn, child, and adolescent health (SRMNCAH) services (1). Disruptions included maternity staff being diverted and hospitals being entirely dedicated for COVID-19 patients, thus reducing facility-based births and limiting access for ANC and PNC; personal financial barriers to access services due to interruption in employment and stressed financial and banking services; physical barriers to access of services due to movement restriction and non-availability of transport during lockdowns; and fears and social stigma surrounding COVID-19 (1, 5-7). It was estimated that health care utilisation has decreased by approximately one-third during the pandemic (8).

Telehealth is an important tool in the provision of health care services, and one strategy that can be employed to address the low service utilisation during the pandemic. However, the study demonstrated that the use of telehealth for the provision of maternal and newborn health care was challenging. Some of the challenges experienced by countries, include the lack of access to technology, impact on ability to build trusting caregiver-client relationships, and, potential for lower quality care leading to further health inequalities (6). This study explored the experiences of key health system stakeholders, maternal and newborn health care providers and childbearing women when adapting to service provision and access changes in response to the COVID-19 pandemic in parts of Bangladesh and Laos PDR. The service provision and access change was the sudden pivot from face-to-face to remote (telehealth) care provision for routine ANC and PNC supported by guidance from the United Nations Population Fund (UNFPA) (7, 8) a key agency supporting the provision of maternity care in both countries. The aim of this study was to explore the experiences of key stakeholders, healthcare providers and childbearing women regarding the implementation of telehealth for routine ANC and PNC during the COVID-19 pandemic in selected regions of Bangladesh and the Lao People’s Democratic Republic (PDR). The research also sought to examine the barriers and enablers of adaptation and implementation of telehealth to manage the impact of COVID-19 on the delivery of ANC and PNC.

## METHODS

In selected regions of Bangladesh and Lao PDR, a qualitative examination of experiences with maternity service provision changes was undertaken. Data were collected through semi-structured interviews with health service leaders and providers, and childbearing women who organised, provided, or were the recipients of telehealth during the COVID-19 pandemic. A framework analysis process was used. Human research ethical approval was received through Alfred Health in Victoria, Australia (HREC Approval Number: 30/21) and through available processes in Bangladesh (CIPRB/ERC/2021/02) and Lao PDR (Submission ID 2021-24) prior to commencement of the research.

### Participants

The study participants included childbearing women who received telehealth services for ANC and PNC; health service providers, the majority of whom were midwives, but also included obstetric and general doctors, nurses, and medical assistants. Key stakeholders included health service managers, Ministry of Health representatives, district health officers and United Nations (UN) staff involved in supporting maternity service provision. An official government letter was issued to each of the facilities about interviewing participants of the study.

Participants were purposively recruited, with key stakeholders being identified by UNFPA, a key agency supporting Ministries of Health in the provision of maternity care in both countries. Health care providers involved in the roll-out of telehealth services were identified by health service managers and invited to participate either by email or phone by the research coordinators in each country. Women who received telehealth for one or more ANC and PNC contact visits were identified from health service records and invited through telephone contact.

Following identification and contact by the research team, participants were provided with the Participant Information Sheet and Consent Form, and given the opportunity to ask further questions about the study. They were reassured that their involvement was voluntary, their contributions would be kept confidential, and participation would not impact on employment or service provision.

### Data Collection

Between October and November 2021, data were collected through semi-structured interviews undertaken at a time and place convenient to the participant. There were 67 interviews conducted in Bangladesh and 34 in Lao PDR (Table 1). The interviews were conducted in English, where the participant identified English as an option, or in the participants’ language by a native speaking interviewer. All interviews were audio-recorded and translated into English where required, and transcribed verbatim.

**Table 1:**
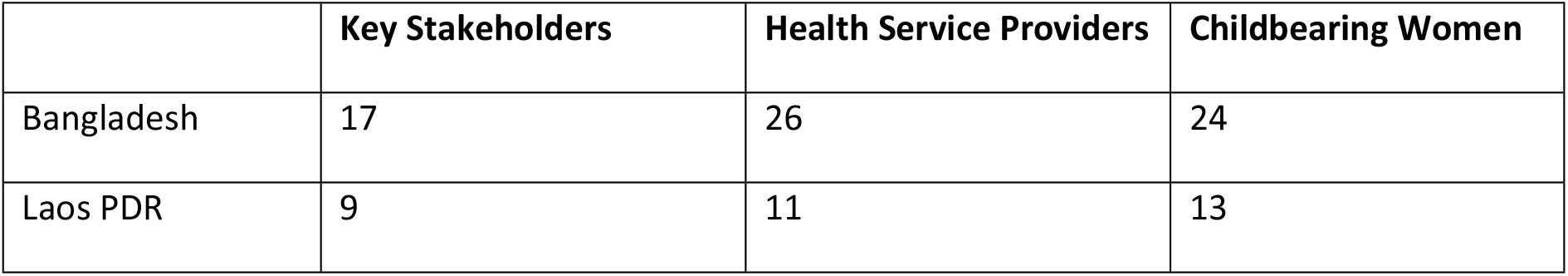
Number of interviews by participant group and country.

|  | **Key Stakeholders** | **Health Service Providers** | **Childbearing Women** |
| --- | --- | --- | --- |
| Bangladesh | 17 | 26 | 24 |
| Laos PDR | 9 | 11 | 13 |

Interviews with key stakeholders were no longer than 60 minutes and were conducted remotely through the online platform Zoom by one of the research team (ST) or a local UNFPA representative. Interviews with the health service providers and women were conducted face-to-face or by telephone using the set of semi-structured questions designed for each cohort. The questions for the semi-structured interviews were developed by the research team, discussed, and refined through the development of the research protocol and ethics application and data collectors received training in use of the tools. Individuals were interviewed in their ‘personal’ capacity; they provided personal opinions to interview questions rather than giving views as employees of any institution.

Interviews with key stakeholders focused on the health systems responses in relation to maternal and newborn care. Participants were asked to identify changes in health service provision, discuss barriers, enablers, and challenges to implementing changes and whether the changes might be sustained beyond the pandemic. Health provider questions focused on whether planning and support for the service changes was provided and the challenges associated with delivering care through telehealth. Childbearing women were asked to identify the impact COVID-19 had on their experience of pregnancy, birth, and early parenting and what they thought about receiving ANC and PNF services through telehealth.

### Data Analysis

Qualitative data were analysed by using framework analysis (9, 10). Framework analysis is a systematic and stepwise approach to qualitative data analysis and has been used in various contexts of healthcare disciplines, including maternity care. Framework analysis offers a pragmatic approach suitable for health care policy and practice, and permits a structured but flexible data analysis process (11). An advantage of framework analysis is the inclusion of a priori coding (based on the literature) and the generation of iterative codes from data. This made it possible to focus the findings on the specific project aims, leading to meaningful conclusions, and recommendations for future system improvements. A further advantage is the ability to manage data from different sources; as this study includes different groups of participants from different countries, so commonality and diversity of views can be systematically identified. The steps used in framework analysis included the setting of clear aims and objectives for the research, initial and ongoing familiarisation with data, development of codes using NVivo Software, including indexing and charting, summarising/interpretation of data, and the development of themes.

To ensure accurate interpretation of the data, quality control measures were put in place. Data were systematically coded by one researcher (ST), initial coding was reviewed by a second researcher (RS) and developed codes and themes were discussed and further refined by team members (ST, RS, CH, C B-K). Once the preliminary analysis was completed, consultation sessions via webinar were held with all researchers, some participants and with the UNFPA country offices.

We present quotes to illustrate the findings. We give the country(Bang = Banladesh; Lao = Lao PDR) and the participant group (S = stakeholder; HSP = health service provider; W = childbearing woman).

## RESULTS

Three common themes were identified across all data, including quality and access to maternal and newborn health services during the pandemic; adapting innovative approaches to address the needs during the pandemic; utilising telehealth for maternity services, and enablers and barriers to telehealth (Table 2). The summary of findings are presented as per the identified participant groupings of key stakeholders involved in the introduction and oversight of telehealth services for ANC and PNC contacts; health service providers who used remote contacts for care provision; and childbearing women who experienced remote ANC and PNC contacts.

**Table 2:** Themes identified by each group and across all data.

| Participants | Themes by participant group | Common themes across all groups |
| --- | --- | --- |
| **Key stakeholders** | ▪ Supports and resources<br>▪ Workforce preparedness<br>▪ Developing, adapting and utilising guidelines and policies during the pandemic<br>▪ Partnership and collaborations<br>▪ Enablers and barriers of implementing telehealth | ▪ Quality and access to maternal and newborn health services during the pandemic,<br>▪ Adapting innovative approaches to address the needs during the pandemic,<br>▪ Utilising telehealth for maternity services and its enablers and barriers |
| **Providers** | ▪ The impact of the pandemic on maternity services<br>▪ Benefits and challenges using telehealth for maternity care<br>▪ Need for training and supportive supervision of midwives<br>▪ Lack of community awareness about telehealth |  |
| **Women** | ▪ Challenges in access to mobile phone and technology<br>▪ Satisfaction and dissatisfaction with remote care services |  |

### Experiences of key stakeholders

The impact of COVID-19 on maternal and newborn health services was significant with a general lack of preparedness to respond to a crisis such as the pandemic. There was a lack of budget to respond to the increased burden on health facilities and this included funds for purchasing essential personal protective equipment (PPE) and other supplies needed to keep maternity staff and clients safe. Key stakeholders identified the pervasive environment of fear that existed in services, among service providers and for women and communities. In part, this fear and lack of strong infrastructure such as access to transport in lockdowns facility closures, led to low service utilisation.

Stakeholders had to respond urgently to the evolving situation, which necessitated them trying to gather and combine information to best inform country-specific needs. The following quote demonstrates the urgency of the situation:

> *… we tried to pull together information from UNFPA and from WHO [World Health Organization] and from other sources from the internet where we could get access in order to integrate it to fit the situation in Lao. (Lao-S)*

There was a need for inter-agency and government collaboration to protect the continuation of essential SRH services. This stakeholder identified a number of issues for consideration:

> *… what would they do if somebody came in labour, WHO was already addressing issues but mostly with the focus on just COVID patients who needed COVID care, but not what you do to keep the maternity patients safe and what do you do with a maternity patient who has got COVID and she is giving birth right that minute. (Lao S)*

Restructuring the physical health service space was undertaken in response to the pandemic and this included allocating separate spaces and entrances for suspected COVID-19 cases. The development, timely distribution, and implementation of national, regional, and global guidance was seen as a key action to guide service providers and protect maternal and newborn health (MNH) services. The provision of UNFPA Technical Guidance assisted the implementation of telehealth for ANC and PNC service provision. The UNFPA Technical Briefs (7, 8) were used to support Government and other NGO responses to the changes needed to ensure continued and safe maternity service provision, as evidenced in the quotes below:

> *… we shared the UNFPA guide at a very high level, we made sure that it was the basis for the national guidelines, and we also shared it in our humanitarian response, we shared it with WHO, we got the plans around how to support the health facilities, both in development context and in our humanitarian context were all guided by the original guidelines. (Bang-S)*

Stakeholders appreciated the clarity of the UNFPA Technical Briefs and the timeliness of the provision of the guidance documents.

> *UNFPA regularly provided us guidelines and a clear-cut guidelines, clear instructions. There is no ambiguity of the information*. *(Bang-S)*

Other resources that were found to be beneficial included the on-line training provided with the UNFPA Technical Briefs. Stakeholders indicated the training was beneficial and that having the training in electronic form that could be shared widely. One stakeholder stated:

> *There had been a webinar that went over a virtual antenatal visit and a lot of the midwives joined that webinar, and it was recorded so we shared it with people, and it was also shared in our training, but they were also given a checklist of what they would cover for antenatal care and what they would cover for postnatal care. (Bang-S)*

When asked about the benefits and challenges of introducing telehealth for ANC and PNC contacts, the majority of stakeholders could see the benefits but also were cognizant of the challenges of rapid implementation. For example:

> *… they [the government services] have to be ready and they need technical support. If you just hand it [telehealth] to the government, later it’ll be seen that it’s not possible. Not all over the country. (Lao-S)*

> *If it is to be done officially, it has to be built on a foundation. Then, it’ll exist or merge with the government’s program as a program. (Lao-S)*

It was evident that there were benefits for women who were offered telehealth services. One stakeholder explained this by saying:

> *It should be done everywhere because many mothers cannot come to the hospital, there are many problems at home, in that case they can take the service for the baby or mother. They are very happy to get the mobile number because if they can talk on the mobile, the problem is less, they get some solution or advice on what to do. (Bang-S)*

Another stakeholder identified benefits for service providers and care coverage stating:

> *One of the off-shoots is also the increase in postnatal care, because previous to this, you know providers focused a lot on the antenatal piece, but postnatal care had been always on the backburner, but because of the ease of getting to the clients, just to phone and check … they found it easier to follow up, and having really strong links with the village health volunteers helped them to connect with the clients better. (Lao-S)*

Stakeholders shared some of the innovations introduced to support service provision. These included utilising community volunteers to support women in accessing telehealth services, the introduction of outreach services and home visiting. Another innovation was a more efficient process for record keeping and documentation of ANC services. One stakeholder explained the changes in record keeping as follows:

> *The midwives had to make an appointment book, they had to see the woman and then schedule her and then write it down for themselves, because they weren’t telling her [the woman] to call them, they were going to call her, so they had to make an appointment book and call her, which had never been done in the public system. (Bang-S)*

While there are similarities in the views of stakeholders from both countries there are some issues that mainly raised by Bangladesh’s key stakeholders, such as gender barriers and its impact on accessing technology and use of telehealth and community involvement, as illustrated below:

> *…w*h*en midwife calls that mother, they are busy. It’s one of the challenges. Some mothers have no mobile phone and instead they provided their husband’s number and therefore the midwife could not provide information because the mobile phone is their [husbands] property. Some mothers are not allowed to talk, instead their mother-in-law was responding to those calls. (Bang-S)*

Overall, the stakeholders described the significant impact of COVID-19 on the provision of maternal and newborn health services but also identified enablers and barriers to implementation that need to be considered going forward. When discussing planning for the future, there needs to be government stewardship, community awareness raising and increased funding to enable scale-up of telehealth services.

### The experiences of health service providers

The health service providers, the majority of whom were midwives, reported considerable impacts from the pandemic and challenges providing telehealth services. COVID-19 created an environment of fear, leading to low utilisation of services, and service shutdowns. Health service providers recognised that the health system was not prepared for a pandemic response. Key issues also included the same issues especially funding to have access to PPE and the inability to keep suspected COVID-19 positive patients separate from other health care users.

A lack of awareness of the community about pandemics created additional challenges for health care providers to continue to provide essential services. One health provider explained how they worked with communities to overcome challenges:

> *The villagers have not yet understood [the pandemic], they are scared to come for services… we did the health education combined with VHV [Village Health Visitor] and head of the village (Lao-HSP)*.

Although health service providers and leaders were aware of the presence of guidance documents, most providers were not aware of detailed guidance, including the UNFPA technical briefs to support the initiation and continuation of telehealth for ANC and PNC. They followed the broader guidelines such as social distancing, use of PPE and hand hygiene as communicated by the head of the facility and the Government. Awareness of the technical guidance increased as evidenced by this quote:

> *I’ve never used UNFPA Technical Briefs before. But in the last training Technical Briefs have been mentioned. So, I will start it now (Bang-HSP)*

Health service providers reported that access to the technology (mobile devices) was not equitable across the population. As described earlier, often they were given numbers of partners or male relatives which made contacting the woman more challenging. Many women did not have access to technology, technology support and insufficient stability of telehealth using mobile phones, as explained:

> *If we call the mother, it can be seen that her brother is receiving or her husband is receiving, or she is in her father’s home (Bang-HSP)*

Providers also reported unstable communication networks, as evidenced here:

> *There is only a problem with the telephone signal, some places do not have a signal, most of the area in [Region] and [Region] have no signal at all, which makes it very difficult to contact pregnant women (Lao-HSP)*

The providers, however, were also able to identify how telehealth made service provision easier in an environment of fear and panic, saying:

> *Suddenly, the number of patients was reduced during corona [COVID-19] as everyone was panicked. But we have to provide services to mothers, and then we did it through the mobile. (Bang-HPS)*

For some, the provision of telehealth services increased the understanding of the role of the midwife and that the women appeared satisfied with this form of care by a midwife:

> *… mothers did not know what a midwife is or what are they doing, but through this phone call, midwives are getting a lot of recognition in Bangladesh. When I leave them with antenatal care, then when I give them a phone call. They are very happy when we give them phone calls, [we get] their positive feedback. (Bang-HSP)*

Health service providers identified supportive actions that assisted in the implementation of telehealth services such as the provision of training, supportive supervision and the technology required to provide the remote contacts. One participant said:

> *But it [telehealth] needs to be made usable. Necessary materials, necessary manpower and equipment support must be provided. Manpower is the main support. Whether there is that support? It takes more materials to give that support. Given these supports, I think it’s definitely a good initiative. I’m in favour of telemedicine. But with all the support. (Bang-HSP)*

When identifying future needs for continued or broader implementation, the health service providers identified the need for increased community awareness, better funding and increased access to technology required to provide the telehealth service.

### Experiences of childbearing women

Most childbearing women interviewed discussed the challenges of accessing to technology and telehealth services. Most women received one or two remote care contacts via phone and the majority of those who utilised telehealth appreciated the service. This was more evident for those women who lived far from their health post. Many women reported limited access to their own mobile devices, and many women relied on their husbands’ mobile to receive consultations. All women had concerns about being infected with COVID-19 in the health facility. Due to their fear of the pandemic and crowds in the health facilities, women avoided going to the health facility. However, most also wanted the option of face-to-face contact. Some reasons for preferring face-to-face contacts are evidenced in the following quotes:

> *It would be better to go to the hospital, because when we come, we will know the baby is good or not. I was happy when they called to ask about the symptoms, but it would be better if I could come and check myself for confidence. (Lao-W)*

> *I don’t like the mobile phone at all, other may like it. If I go to the hospital then they first check my blood pressure, it’s very positive for a pregnant mother. Second thing is, she checks my weight, my baby’s response, but I don’t get these service over the phone - for these reasons I visit there by myself. Many times they requested me to share those problems over the mobile phone but I replied, if I have the time then I must come. (Bang-W)*

Most women reported that they received one to two remote care contacts via mobile phone during their pregnancy. Some allocated to receive remote contacts did not receive a call throughout the pregnancy. This woman reports postpartum only contacts:

> *They called for the first time, when the child was 28 days old. Then, rang once when the child was one and a half months old. (Bang-W)*

There were positive statements about the telehealth services in both countries:

> *I liked that they called me. Because they are taking care of me, taking news, not having to go to the hospital, giving advice about my health. (Lao-W)*

> *it would be good to give this service to everyone. Everyone needs to know that it feels good to get advice at this time. If I get good service, I will definitely tell others about this service. (Bang-W)*

Some women believe the remote care is a good as interim solution during the pandemic however in long term they prefer face to face services.

> *Face-to-face treatment is also a plus. Therefore, in this COVID-19 situation, the online system is the most convenient. (Lao-W)*

As presented, feedback on their satisfaction with the use of telehealth to provide ANC and PNC services was mixed and this feedback should be considered when planning post-pandemic services.

### Lessons Learned

All participants provided valuable information regarding the challenges, enablers and opportunities raised by the need to pivot routine services to remote contacts in the COVID-19 country response. More time to put robust systems in place for these changes would have been beneficial. However, many lessons can be learned to inform future care provision opportunities. Figure 1 provides a summary of the data in relation to managing the current COVID-19 situation, innovations and approaches that support the implementation of telehealth services for the effective use of telehealth services and using the current pandemic services adjustments as an opportunity for change in future care provision and pandemic and other disaster preparedness.

**Figure 1:**
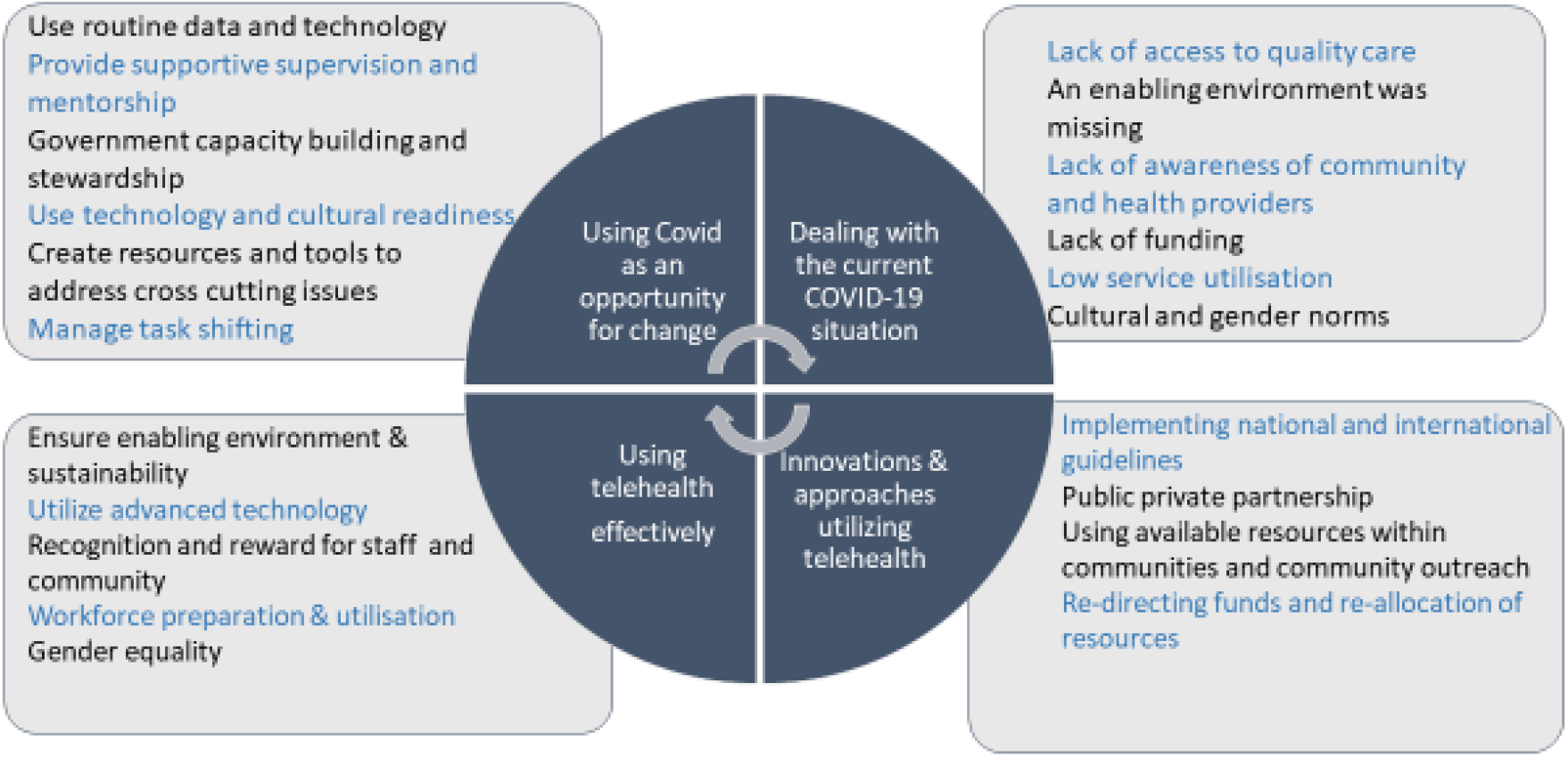
Managing telehealth implementation in COVID-19 context.

## DISCUSSION

This study analysed data from more than 70 interviews with health service managers and leaders, healthcare service providers and childbearing women to explore the impact of the pivot to telehealth service provision for ANC and PNC during the COVID-19 pandemic. Whilst all participants were able to identify benefits of the use of telehealth for the provision of care, many also raised concerns related to equitable access, acceptability, and long-term sustainability.

The COVID-19 pandemic health system challenges drove the rapid initiation, or scale-up, of telehealth services in all-income level countries (12). Telehealth was seen as a solution to ensuring the provision of care continued during the times where health systems were under pressure due to the number of people with COVID-19 requiring hospitalisation and care (13). In addition to increased hospitalisations, the pandemic saw non-COVID related service utilisation decrease as communities were locked down and fearful to attend health services due to the risk of COVID-19 transmission (14, 15).

The evidence identifying and measuring the impact of COVID-19 on health systems, including the provision of maternal newborn care, continues. Concern regarding the impact of the pandemic on maternal and newborn mortality was identified early in the pandemic, and the need to continue to provide vital pregnancy and childbearing services was recognised (3, 16). Rapid reports and anecdotal evidence identified issues such as the cessation of routine ANC and the diversion of funding from maternity care to the acute COVID-19 response (4). Our research identified similar issues with both health service managers and leaders, and health care providers identifying a lack of preparedness, a lack of funding, a lack of PPE which all contributed to low service utilisation and loss of some essential maternal child health services. Fortunately, it was recognised that the negative impact of COVID-19 on routine services needed to be urgently addressed and a number of protective initiatives, including the issue of global guidance and the scale-up of telehealth services, were implemented in Bangladesh and Lao PDR.

Health service leaders in both Bangladesh and Lao PDR initiated actions to protect maternal and newborn health services during the early days of the pandemic. There was recognition of the need for interagency and government collaboration to protect essential services. The WHO Global Action Plan for Healthy Lives report identifies that where multi-lateral partners collaborated closely the impact of COVID-19 on essential health services was reduced (17). The need for actions to sustain service provision changes was a key theme of both health leaders and health service providers in our study who saw the opportunity to build on the combined response to the pandemic and integrate some of the innovations into routine practice. The ability to learn from mistakes and successes in the delivery of universal healthcare during and post-pandemic has been identified as crucial to the continued recovery and strengthening of health systems (2, 17-19).

A key success in the implementation of telehealth services in response to the impact of COVID-19 in Bangladesh and Laos PDR was the timely provision of technical guidance documents (7, 8, 16) designed to support the transition from face-to-face to remote contacts for routine ANC and PNC. Health leaders and providers identified access to online training alongside the provision of guidance allowed for broad dissemination of information and supported the health service providers in the swift service provision changes. Limited training on both care provision, and use of telehealth technology were barriers to implementation. Generally, the uptake of innovation, and in particular the introduction of technology, in health service provision changes are notorious for being slow or difficult to implement (20, 21).

Our findings identified enablers and barriers to implementation requiring leadership and the provision of guidance and support; interagency and government collaboration on management of the program; effective communication regarding dissemination of guidance documents and funding for implementation (22). The urgency caused by the pandemic did not allow for a timely and controlled design and implementation, nor for extensive stakeholder engagement. There was recognition that not all enablers could be addressed prior to implementation. However, when sustainability and future needs were identified, enablers such as stakeholder engagement in the form of increasing community awareness of the service provision change, and further dedicated training and resources were discussed. Similar studies in the Asian Region identified that while there has been increased utilisation of telehealth during the pandemic there is now urgent need to evaluate and potentially re-design scale-up measures to incorporate wider stakeholder engagement, improvements in integrated technology systems and capacity building in the delivery of effective care remotely (23, 24).

Whilst healthcare providers acknowledged the need for remote service provision due to the impact of the pandemic, many felt that face-to-face consultations were more acceptable as there are many routine activities that cannot be attended to over the phone. Telehealth can be viewed as more convenient for staff and consumers, but the extent and reach of care to those who have trouble accessing technology may pose an additional challenge. Many aspects of maternity clinical care are not feasible remotely and cannot be done via mobile phone and would require advanced technology and new skillsets for providers. Furthermore, shifting the community culture from face-to-face care to remote care requires extensive community awareness and campaign (25). Community awareness about the benefits of remote care is critical to the successful implementation of telehealth (23)

In our study, both the childbearing women and healthcare providers raised the issue of access to technology as a barrier to successful implementation of telehealth. This is not dissimilar to other findings where health providers were concerned regarding the quality of care and the risk of increasing inequalities in healthcare access due to a lack of available technology (6, 26). Much of the evidence discusses the need for caution in sustaining telehealth services introduced during the pandemic without further evaluation on acceptability for both healthcare providers and service users, accessibility, and quality of care (6, 13, 23, 26, 27).

The women in our study identified a level of fear regarding accessing routine maternity services at hospitals and health centers and were generally appreciative of the opportunity to have some routine care provided remotely. Fear of contracting COVID-19 from health services lead a broad decline in service utilisation, including in maternal and child healthcare (1-3, 14, 28). Even though the fear of contracting COVID-19 through attendance at health services was identified, many women reported that although telehealth services were satisfactory and convenient, they continued to want face-to-face visits. This preference for face-to-face healthcare provision is widespread in other studies and, as such, caution is required in post-pandemic planning for scale-up of telehealth services (6, 21, 29, 30).

### Limitations

The findings of this study add to existing and emerging evidence regarding the acceptability of telehealth services in the provision of maternity care. Limitations of our study include selection bias as participants were purposively recruited through identification of participant requirements by senior health leaders in each country.

A major limitation was the quality of interview data from the childbearing participant group. Due to language requirements, it was necessary to use interviewers who were not expert researchers and there was a lack of probing applied to each question and response. This is reflected in the limited presentation of findings for childbearing women. In addition, the use of translation for transcribing may have caused a lack of nuance in understanding intent.

Generalizability of the findings may be impacted by the definition of telehealth being by other researchers. However, within the region where the study was conducted, telehealth has been offered through voice-only devices due to limitation in connectivity and access to technology.

### Implications for implementation and practice

The COVID-19 pandemic necessitated the rapid implementation of telehealth services to ensure continued access to and provision of essential healthcare services in many countries. Rapid implementation of any system of care can be challenging. It is now vital that systems of care introduced as a response to COVID-19 disruptions to health service access and provision are fully evaluated prior to general adoption or scale-up. Research on the impact of implementation of telehealth services for selected ANC/PNC contacts needs to consider health systems, health service providers and the women and families who access these services. Further research with women regarding accessibility, acceptability and quality of care must be undertaken prior to any permanent move to the use of telehealth for for selected ANC and PNC contacs.

## CONCLUSION

This study explored the swift implementation of telehealth for selected ANC/PNC contacts in low-resource settings during the COVID-19 pandemic. The findings provide future direction for the expansion of the use of telehealth in maternity care in low resource settings. Strategic planning for collaboration and partnership with the private sector and Ministry of Health was seen as a strategy to enhance efficacy and accessibility to telehealth. Telehealth supported the delivery of childbearing women’s health needs during the pandemic and could potentially be continued or scaled up in the post-pandemic recovery.

A structured framework for training and support of health providers on digital literacy and the advancement of innovative technologies compatible with local culture may address some of the challenges and inequalities in implementation in low-resource settings. Adequate supportive supervision is needed to promote the provision of quality care. Midwives need to be prepared and supported to utilise telehealth effectively. The equity of access to technology for all women should be at the forefront of sustained or continued implementation of telehealth programs.

## Data Availability

Data can be accessed via Burnet Institute upon request.

## Acknowledgements

Thank you to the Ministry of Health in Bangladesh and Lao PDR, the resaerch teams and all the individuals who were interviewed for this study.

